# *APOE ε*4 carriage associates with improved myocardial performance in older age

**DOI:** 10.1101/2022.07.20.22277846

**Authors:** Constantin-Cristian Topriceanu, Mit Shah, Matthew Webber, Fiona Chan, James C Moon, Marcus Richards, Nishi Chaturvedi, Alun D. Hughes, Jonathan Schott, Declan P. O’Regan, Gabriella Captur

## Abstract

**Introduction:** Although *APOE ε*4 allele carriage confers a risk of coronary disease, its persistence in human populations might be explained by certain survival advantages (antagonistic pleiotropy).

**Hypothesis:** Combining data from three British cohorts–1946 National Survey of Health and Development (NSHD), Southall and Brent Revised (SABRE) and UK Biobank–we explored whether *APOE ε*4 carriage associates with beneficial or unfavorable left ventricular (LV) structural and functional parameters by echocardiography and cardiovascular magnetic resonance (CMR) in older age.

**Methods:** Based on the presence of *APOE ε*4, genotypes were divided into: *APOE ε*4 (ε2ε4, ε3ε4, *ε*4*ε*4) and non-*APOE ε*4 carriers. Echocardiographic data included: LV ejection fraction, E/e’, systolic and diastolic posterior wall and interventricular septal thickness (LVPWT_s/d_, IVS_s/d_), LV mass and the ratio of the LV stroke volume to the LV myocardial volume called myocardial contraction fraction (MCF). CMR data additionally included longitudinal and radial peak diastolic strain rates (PDSR). Generalized linear models explored associations between *APOE ε*4 genotypes as exposures and echocardiographic/CMR biomarkers as outcomes. As APOE genotype is a genetic instrumental variable (unconfounded), Model 1 was unadjusted; Model 2 was adjusted for factors associated with the outcome (age, sex, and socio-economic position) to yield more precise estimates; and subsequent models were individually adjusted for mediators (body mass index, cardiovascular disease [CVD], high cholesterol and hypertension) to explore mechanistic pathways.

**Results:** 35,568 participants were included. Compared to the non-*APOE ε*4 group, *APOE ε*4 carriers had similar cardiac echocardiographic phenotypes in terms of LV EF, E/e’, LVPWT_s/d_, IVS_s/d_ and LV mass but had a 4% higher MCF (95% confidence interval [CI]: 1–7%, *p*=0.016) which persisted in Model 2 (95% CI 1–7%, *p*=0.008) but was attenuated to 3% after adjustment for CVD, diabetes and hypertension (all 95% CI 0–6%; all *p*<0.070). This was replicated in UK Biobank using CMR data, where *APOE ε*4 carriers had a 1% higher MCF (95% CI 0-1%, *p*=0.020) which was attenuated only after adjusting for BMI or diabetes.

**Conclusions:** *APOE ε*4 carriage associates with improved myocardial performance in older age resulting in greater LV stroke volume generation per 1 mL of myocardium and better longitudinal strain rates compared to non *APOE ε*4 carriers. This potentially favorable cardiac phenotype adds to the growing number of reported survival advantages attributed to *APOE* ε4 carriage that might collectively explain its persistence in humans.

## INTRODUCTION

*Apolipoprotein ε* (*APOE ε*) mediates the biding of low-density lipoprotein (LDL) to peripheral receptors. Given the existence of two single-nucleotide polymorphisms, namely rs429358 and rs7412, there are three *APOE ε* isoforms coded by the alleles *ε*2, *ε*3 and *ε*4 giving rise to six genotypes namely ε2ε2, ε2ε3, ε2ε4, ε3ε3, ε3ε4 and ε4ε4 with the commonest being ε3ε3^1^. *Apolipoprotein ε*4 is regarded to be a major risk factor for developing Alzheimer’s disease^2^ even from young age, especially in females ^3^. In addition, it may associate with decreased physical performance in older age^4^ and decrease cognitive performance (e.g., verbal episodic memory) in healthy young adults^5^. Yet despite its adverse associations, this ancestral allele has persisted in human populations instead of being replaced by the more recently evolved alleles, *ε*3 and *ε*2^6^ suggesting its carriage might be conferring some survival advantages. Indeed, *APOE ε*4 carriers have been shown to have increased fertility^7,8^, resistance to infections^7^, decreased perinatal and infant mortality^7^, decreased chronic airway obstruction ^9^, fewer arterial aneurysms^9^ and peptic ulcers^9^, less liver disease and slight cognitive advantages^7 10^.

In terms of the cardiovascular system, carriage of *ε*4 (rs429358-cytosine and rs7412-cytosine) has been associated with adverse clinical sequelae including ischaemic heart disease (IHD)^11^, hypertension^12^, diabetes^13^ and high LDL^14^. Moreover, heart function was also suggested to be a mediator in the association between *ApoE ε*4 and gray matter decline^15^. However, to date it remains unclear whether *APOE ε*4 carriage independently associates with a better or worse long-term cardiac phenotype in terms of heart size and function. Using cohort data from the Medical Research Council (MRC) 1946 National Survey of Health and Development (NSHD), Southall And Brent Revised (SABRE) and United Kingdom (UK) Biobank, we explored this association.

## METHODS

### Study population

The MRC NSHD is the world’s longest-running birth cohort with continuous follow-up. In 1946 in Britain, 5,362 individuals (2547 males and 2815 females) born in the same week in March were enrolled. Participants were invited for periodic follow-ups in which health and socio-economic assessments were performed which have been described elsewhere^16^.

The SABRE study is a tri-ethnic cohort of European, South Asian, and African Caribbean participants living in North and West London. Between 1988-1981, participants aged 40-69 years were randomly selected from 5-year age and sex stratified primary care lists (n=4063) and workplaces (n=795). Full details have been described elsewhere^17^.

The UK Biobank is a large prospective cohort study with more than half a million individuals recruited between 2006 and 2010 when study participants were aged 40-69 years old, and features demographic, genetic, health outcome and imaging data for participants. ^18^. Details of subjects’ comorbidities were obtained through self-reported diagnoses and International Classification of Disease (ICD-9 and ICD-10) codes from linked medical records This project was conducted using the UK Biobank (UKBB) resource under application numbers 40616 and 46696.

### Data availability

NSHD data is available from: https://www.nshd.mrc.ac.uk/data, SABRE data is available from https://www.sabrestudy.org/, and UK Biobank data is available from https://www.ukbiobank.ac.uk/.

### Ethical approval

The 2006-2010 NSHD data collection sweep included an in-depth cardiovascular assessment and was granted ethical approval from the Greater Manchester Local Research Ethics Committee and the Scotland Research Ethics Committee^16^ and written informed consent was given by all study participants. Similarly, the SABRE study was granted ethics approval from Ealing, Hounslow and Spelthorne, Parkside, and University College London Research Ethics Committees with all participants giving written consent. Our project was approved by both the SABRE and NSHD committees. UK Biobank’s ethical approval was from the Northwest Multi-centre Research Committee (MRCEC) in 2011, which was renewed in 2016 and then in 2021. All procedures performed were in accordance with the ethical standards of the institutional and/or national research committee and with the 1964 Helsinki declaration and its later amendments or comparable ethical standards.

### Outcomes: Echocardiographic data

In NSHD, when study members were 60-64 years (2006-2010), British-based NSHD participants who had not been lost to follow-up or withdrawn, were invited to attend a clinic-based assessment that included resting transthoracic echocardiography using General Electric (GE) Vivid I machines. The echocardiographic protocol included long and short axis (LAX and SAX), apical 5-, 4-, 3- and 2- chamber, aortic SAX views ^19^. In SABRE, study members were invited between 2008 and 2012 to a clinic visit in which echocardiographic data was acquired using a Phillips iE33 ultrasound machine S5-1 phased array and a X3-1 matrix transducer and analyzed using Philips QLAB software 7,0^17^ in line with the with the American Society of Echocardiography (ASE) guidelines^20^. In both cohorts, echocardiographic data provided left ventricular (LV) ejection fraction (EF), E/e’, systolic and diastolic LV posterior wall and interventricular septal thickness (LVPWTs/d, IVSs/d), LV mass (LVmass). Myocardial contraction fraction (MCF) was calculated as the ratio between stroke volume and myocardial volume. Although indexation to body surface area (BSA), is commonly done in clinical practice, BSA is a poor indexation metric as it creates a bias for overweight individuals^21^. Although indexation to allometric height is a better alternative^21^, indexation might lead to spurious associations, as the exposure might be associated with height/weight rather than with the outcome itself. Therefore, we used unindexed echocardiographic outcomes in all subsequent analyses.

### Outcomes: Cardiovascular magnetic resonance data

Participants in the UK Biobank were randomly invited for a CMR scan on a 1.5 T Siemens Aera scanner from 2014. Briefly, the CMR imaging protocol consisted of three long-axis views and a complete short axis stack of balanced steady state free precession cines^22^. Grey-scale short axis cine stacks were automatically segmented using a deep learning neural network that has optimised for UKBB scan images, with human expert level performance ^23^. The short-axis segmentations underwent post-processing to compute end-systolic, end-diastolic and stroke volumes in both ventricles ^24^. Left ventricular mass (LVM) was computed from left ventricular volume (assuming a density of 1.05 g/ml). Left ventricular wall thickness was computed as the perpendicular radial-line distance between endocardial and epicardial surfaces at end-diastole for each of the 17 myocardial segments as defined by the American Heart Association (AHA)^25^. MCF was derived as above. Thickness of the IVS was calculated as the mean wall thickness of segments 2, 3, 8, 9 and 14, while PWT was taken as the mean of segments 5,6, 11, 12, and 16. To compute longitudinal and radial peak diastolic strain rates, non-rigid image co-registration was performed between successive frames to enable dynamic motion tracking of the heart during the cardiac cycle ^26^. Unindexed CMR metrics were used in all subsequent analyses as discussed above.

### Exposures: *APOE ε* genotype

In NSHD, blood samples were collected at age 53 by a trained research nurse, and DNA was extracted.^27^ Genetic analysis of stored samples took place in in 1999 and 2006-2010. In SABRE, blood samples were collected during baseline studies in 1988-1991 and during follow-up from 2007-2012^17^. Genotyping of rs439358 and rs7412 was conducted at the Exeter University for SABRE and by LGC, Huddleston, UK for NSHD^28^.. Genotyping of UK Biobank participants is detailed elsewhere ^29^, however in brief, genotyping for 488,252 subjects was performed using the UK BiLEVE or UK Biobank Axiom arrays and imputation based on the HaplotypeReference Consortium and UK10K+1000 Genomes panels. Imputation V3 (in GRCh37 coordinates) was used for the current study. Genotypes in their released PLINK-format files were used on the DNANexus platform (https://www.dnanexus.com/). Based on the presence or absence of *APOE ε4*, genotypes were categorically defined as: non-*APOE ε*4 carriers (*ε*2*ε*2, *ε*2*ε*3, *ε*3*ε*3), heterozygous-*APOE ε*4 (ε2ε4 and ε3ε4) or homozygous-*APOE ε*4 (*ε*4*ε*4). Heterozygous-*APOE ε*4 and homozygous-*APOE ε*4 were further grouped into *APOE ε*4 carriers.

### Covariates

Sex was recorded as male or female. The age, weight, and height at the time of the imaging were used to compute the body mass index (BMI) in all 3 cohorts. In NSHD, participants’ socioeconomic position (SEP) was evaluated at the time of echocardiography according to UK Surveys Registrar General’s social class, dichotomized as manual or non-manual. In UK Biobank, we used the Townsend deprivation index scores derived from national data about ownership and unemployment aggregated by postcodes^30^. The presence of cardiovascular disease (CVD), diabetes or high cholesterol was recorded as 1=present or 0=absent.

### Statistics

All analyses were performed in R 4.0 ^31^. For all analyses, a two-tailed *p*-value <0.05 was considered statistically significant.

Distribution of data were assessed on histograms and using Shapiro-Wilk test. Continuous variables are expressed as mean ± 1 standard deviation (SD) or median (interquartile range) as appropriate; categorical variables, as counts and percent.

In the main analysis, we compared non-*APOE ε*4 carriers with *APOE ε*4 carriers. Given the skewed distributions of echocardiographic and CMR data, generalized linear models with gamma distribution and log link were used to investigate the association of *APOE ε4* genotypes as the exposures to predict the continuous echocardiographic and CMR variables as the outcomes. As the longitudinal and radial PDSR also spanned negative values, generalized linear models with Gaussian distribution and identity link were used instead. Being a combination of gene variants, *APOE ε* genotype is expected to be an instrumental variable and therefore unconfounded. Thus, Model 1 was unadjusted. To obtain more precise regression estimates, Model 2 was adjusted for factors associated with the outcome, namely age, sex, and SEP. To explore the mechanistic pathway downstream of *APOE ε* genotype but upstream of the echocardiographic outcomes, subsequent models were adjusted for mediators as follows: Model 3 for BMI; Model 4 for the presence of CVD; Model 5 for diabetes; Model 6 for high cholesterol; and Model 7 for hypertension (**Figure 1**). Model assumptions were verified with regression diagnostics and found to be satisfied.

**Figure 1.**
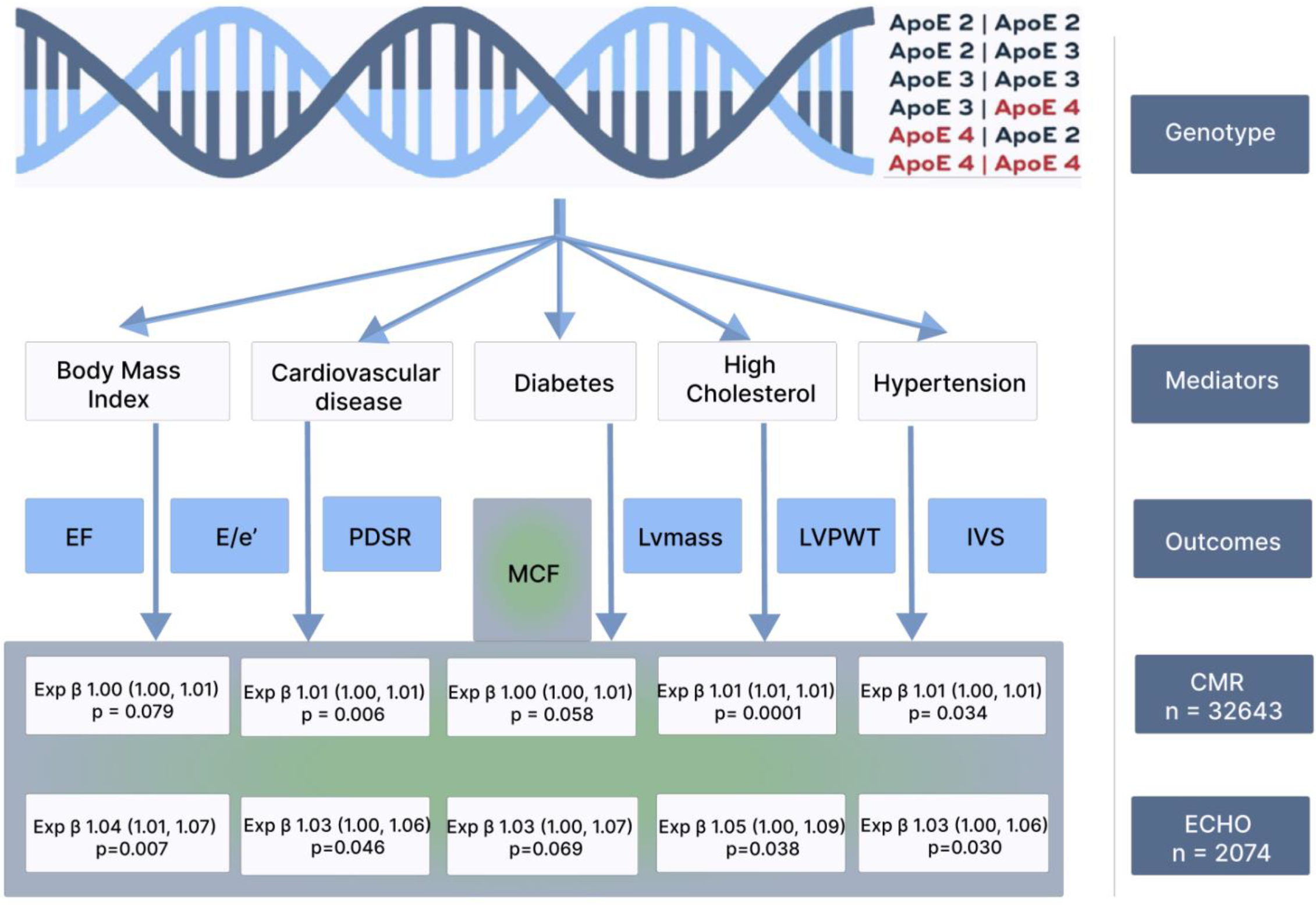
Associations between *APOE ε4* genotypes and echocardiographic and cardiac MRI data in older age. As *APOE ε4* carriers had a higher myocardial contraction fraction, the mechanistic pathways were explored by adjusting the models for mediators (body mass index, cardiovascular disease, diabetes, high cholesterol, and hypertension). *EF, ejection fraction; IVS, interventricular septal thickness; LVmass, left ventricular mass, LVPW left ventricular posterior wall thickness; MCF myocardial contraction fraction; PDSR, longitudinal/radial peak diastolic strain rate*.

For all the models, regression estimates were obtained separately for NSHD, SABRE and UK Biobank (i.e., cohort specific analyses). Since both NSHD and SABRE participants had echocardiography, random-effects meta-analyses were performed across these 2 cohorts. Heterogeneity was evaluated using the Cochran Q test and Higgins I^2^ statistic. Since UK Biobank had CMR data, it was not included in the meta-analysis.

To explore dose responses, *APOE ε4* genotypes were recoded as an ordered category based on the number of *ε4* possessed. Thus, class 0 = *ε2ε2, ε2ε3, ε2ε3*; class 1= *ε2ε4* and *ε3 ε4*; and class 2 = *ε4ε4*. Given the existence of 3 classes, generalized linear models with gamma distribution (or Gaussian distribution for longitudinal and radial PDSR) and orthogonal polynomial contrasts with 2 equally spaced levels (i.e., linear and quadratic) were employed to look for a dose response by *ε4* variants.

As a sensitivity analyses, *APOE ε*4 carriers were split into heterozygous-*APOE ε*4 (*ε*2*ε*4 and *ε*3*ε*4) and homozygous-*APOE ε*4 (*ε*4*ε*4), and all the analyses were replicated as above.

## RESULTS

### Participant characteristics

Participants with available *APOE ε*4 genotype and at least one cardiac imaging metric were included yielding a total of 35568 participants (n=1467 from NSHD, n=1187 from SABRE and n=32972 from UK Biobank). Their characteristics are shown in **Table 1**. In total, there were 816 (2.29%) homozygous-*APOE ε*4, and 9103 (25.59%) heterozygous-*APOE ε*4 individuals with a good agreement between NSHD, SABRE and UK Biobank. SABRE participants were more likely to be males (76.75%), have a higher BMI (median 27 years) or suffer from hypertension (58.98%) compared to NSHD and UK Biobank. On the other hand, UK Biobank participants were least likely to suffer from CVD (6.53%), diabetes (18.64%) or hypertension (27.62%).

**Table 1.**
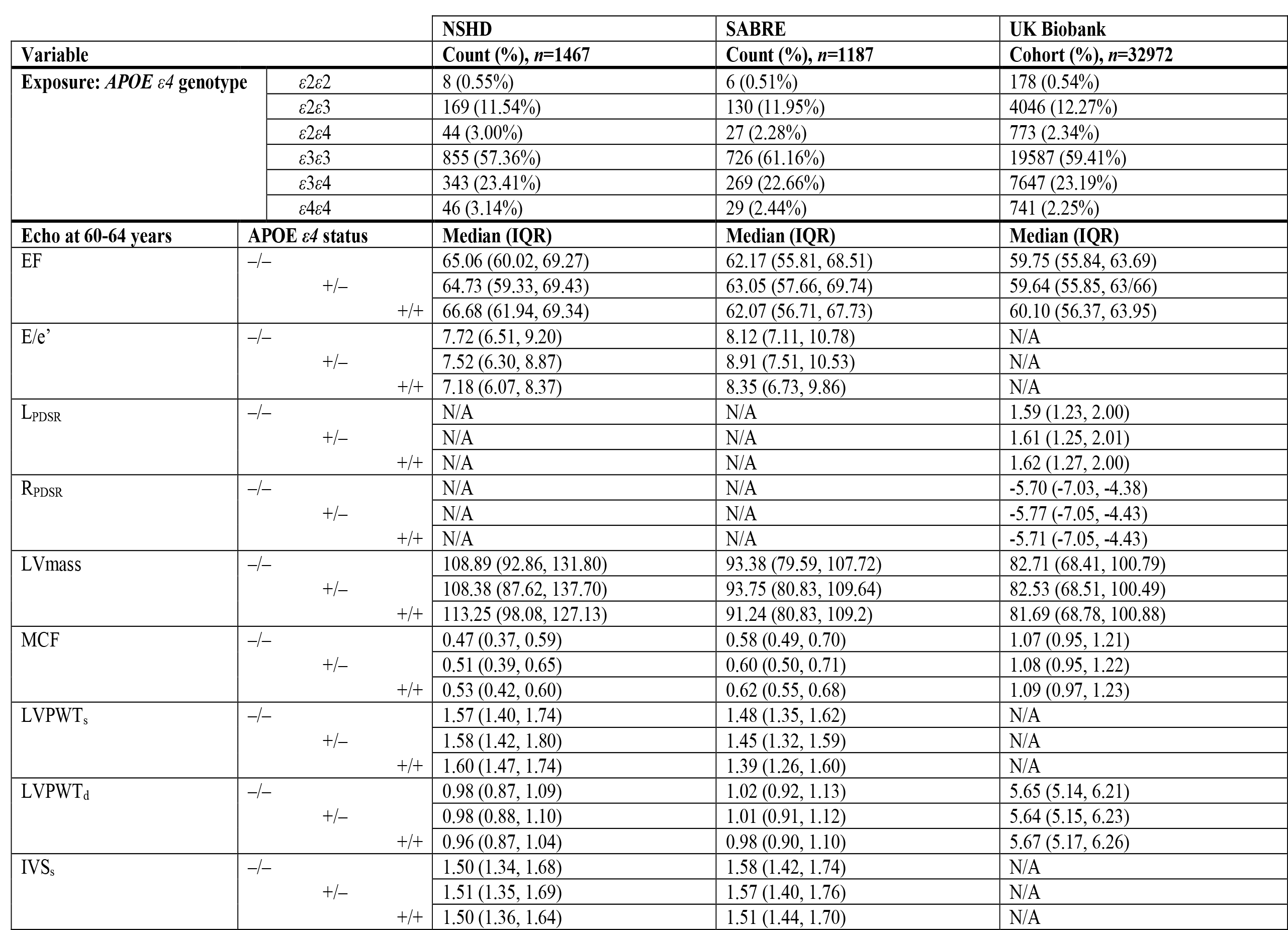
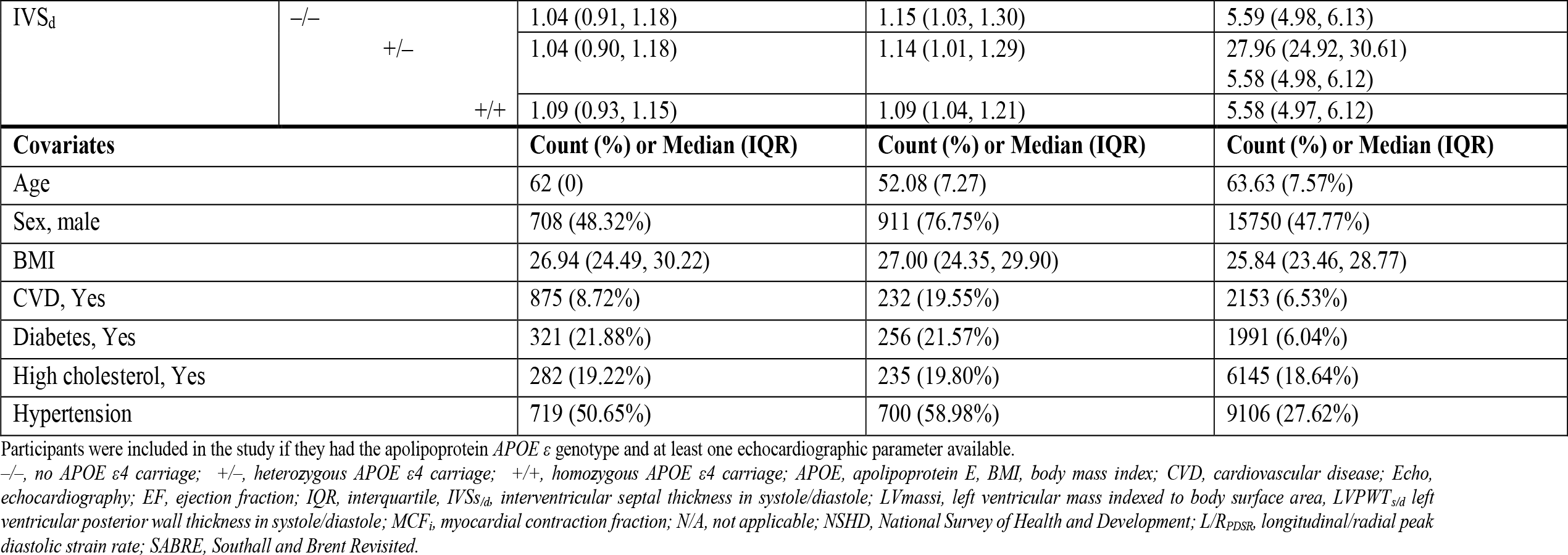
General characteristics of study participants.

### Associations between *APOE ε4* genotypes and echocardiographic data

In NSHD, when compared to the non-*APOE ε*4 group, *APOE ε*4 carriers had a 6% higher MCF (95% confidence interval [CI] 0-12%, *p*=0.050) which persisted after adjusting for sex and SEP (95% CI 0-12%, *p*=0.038) and diabetes (95% CI 0-12%, *p*=0.056), was attenuated to 5% after adjusting for BMI (95% CI 0-11%, *p*=0.064), CVD (95% CI 0-12%, *p*=0.112) and hypertension (95% CI 1-11%, *p*=0.081), and was increased to 8% after adjusting for high cholesterol (95% CI 1-14%, *p*=0.020, **Supplementary Table S1**). Similarly, *APOE ε*4 carriers had a 5% higher LVmass *p*=0.057 which was increased to 6% after adjusting for CVD (*p*=0.040) and hypertension (*p*=0.040), and to 7% after adjusting for diabetes *p*=0.024. No significant associations were found in SABRE (**Supplementary Table S2**).

In the meta-analyses, compared to the non-*APOE ε*4 group, *APOE ε*4 carriers had similar cardiac phenotypes in terms of EF, E/e’, LVPWT_s/d_, IVS_s/d_ and LVmass but had a 4% higher MCF (95% CI 1–7%, *p*=0.016) which persisted after adjustment for sex and SEP (95% CI 1–7%, *p*=0.008) and was attenuated to 3% after adjustment for CVD, diabetes and hypertension (all 95% CI 0–6%, all *p*<0.070, **Table 2, Figure 1**). However, no significant dose response of *APOE ε*4 carriage was found in the association of *APOE ε*4 genotype with MCF (**Table 4, Supplementary Table S3**). In the sensitivity analysis, only heterozygous-*APOE ε*4 carriers had a 4% higher MCF (95% CI 1-7%, *p*=0.016) which persisted after adjusting for sex and SEP (95% CI 1-7%, *p*=0.013), and BMI (95% CI 1-7%, *p*=0.018) but was attenuated to 3% after adjusting for CVD (95% CI 0-6%, *p*=0.043, diabetes (95% 0-7%, *p*=0.060), and hypertension (95% CI 0-6%, *p*=0.028, **Table 5, Supplementary Table S4**).

**Table 2.**
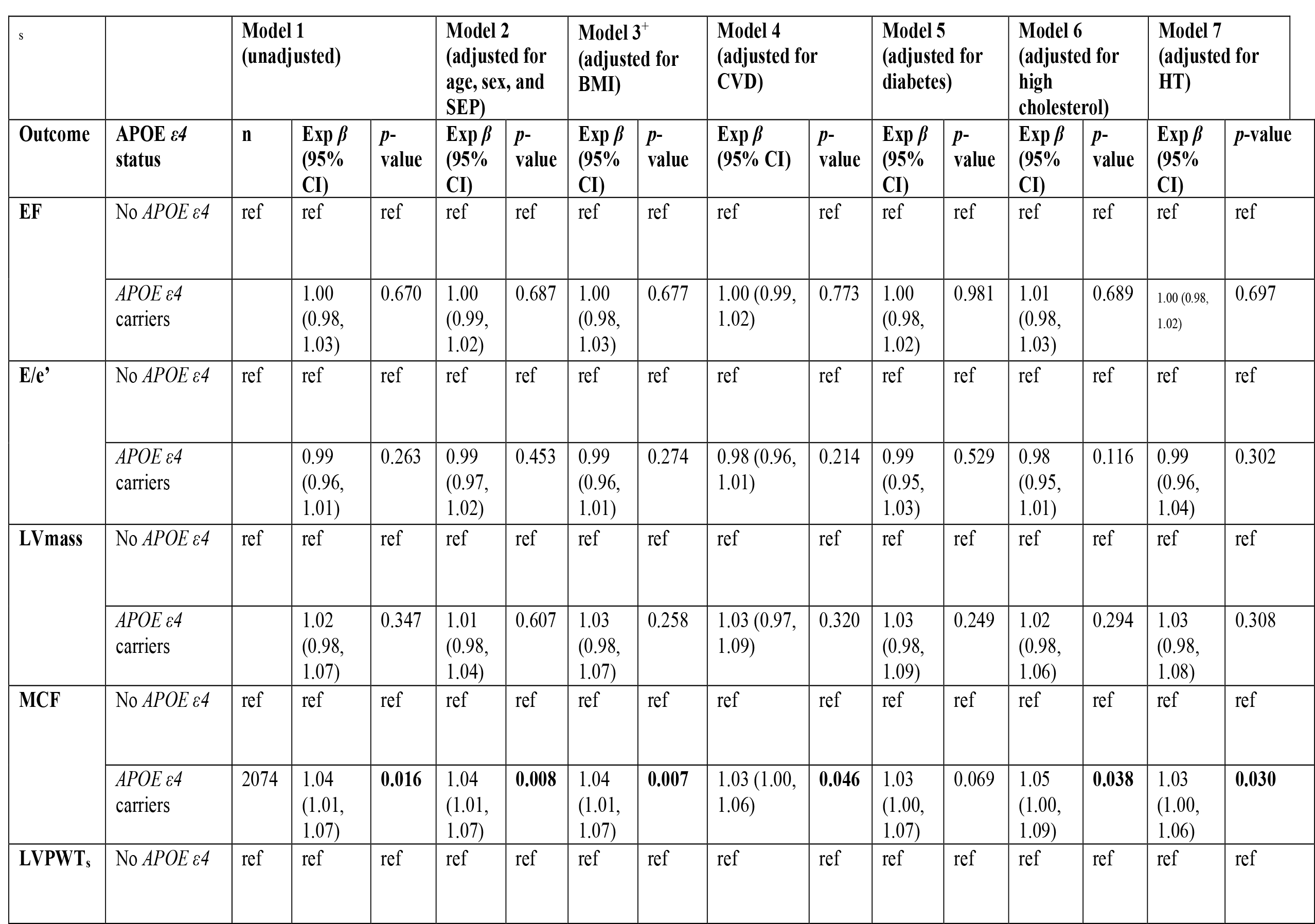
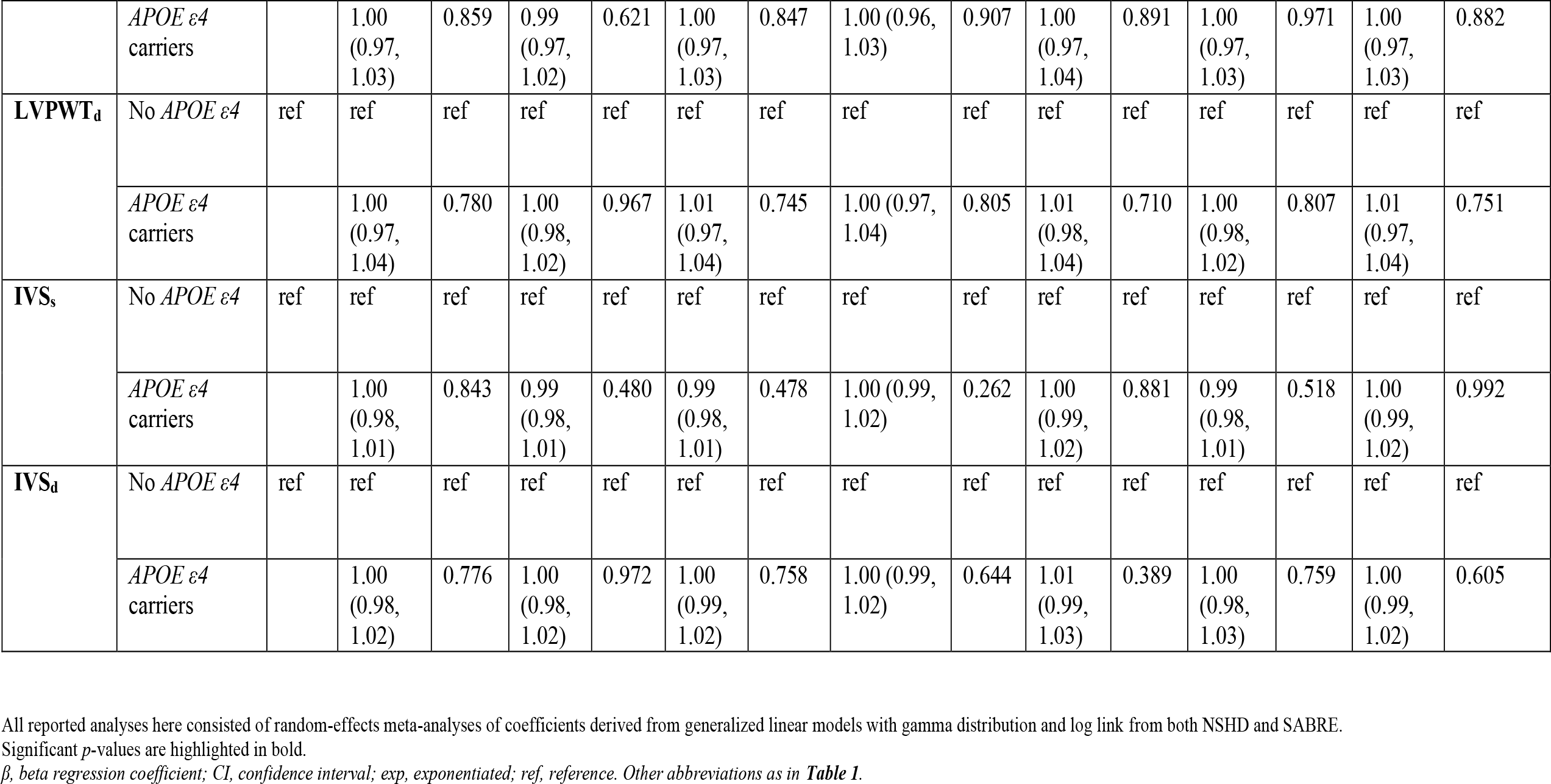
Associations between *APOE ε4* genotypes and echocardiographic data in older age by comparing non-*APOE ε4* (*ε2ε2, ε2ε3, ε2ε3*) with *APOE ε4* (*ε2ε4, ε3ε4 and ε4ε4*) genotypes in the meta-analysis pooling SABRE and NSHD data.

### Associations between *APOE ε4* genotypes and CMR data

In UK Biobank, when compared to the non-*APOE ε*4 group, *APOE ε*4 carriers had a 1% higher MCF 95% (CI 0-1%, *p*=0.020) which persisted after adjusting for age, sex and SEP (Model 2, *p*=0.080), CVD (Model 4, *p*=0.006), high cholesterol (Model 5, *p*=0.0001) and hypertension (Model 7, *p*=0.034) but was attenuated to 0% (95% CI 0-1%) after adjusting for BMI (Model 3, *p*=0.079) or diabetes (*p*=0.058, **Table 3, Figure 1**). There was a dose-response relationship especially when adjusting for CVD in Model 4 (*p*=0.036) and high cholesterol in Model 6 (*p*=0.006, **Table 4**). However, although heterozygous-*APOE ε*4 carriers had a higher MCF, the association was not significant for homozygous-*APOE ε*4 carriers (**Table 5**).

**Table 3.**
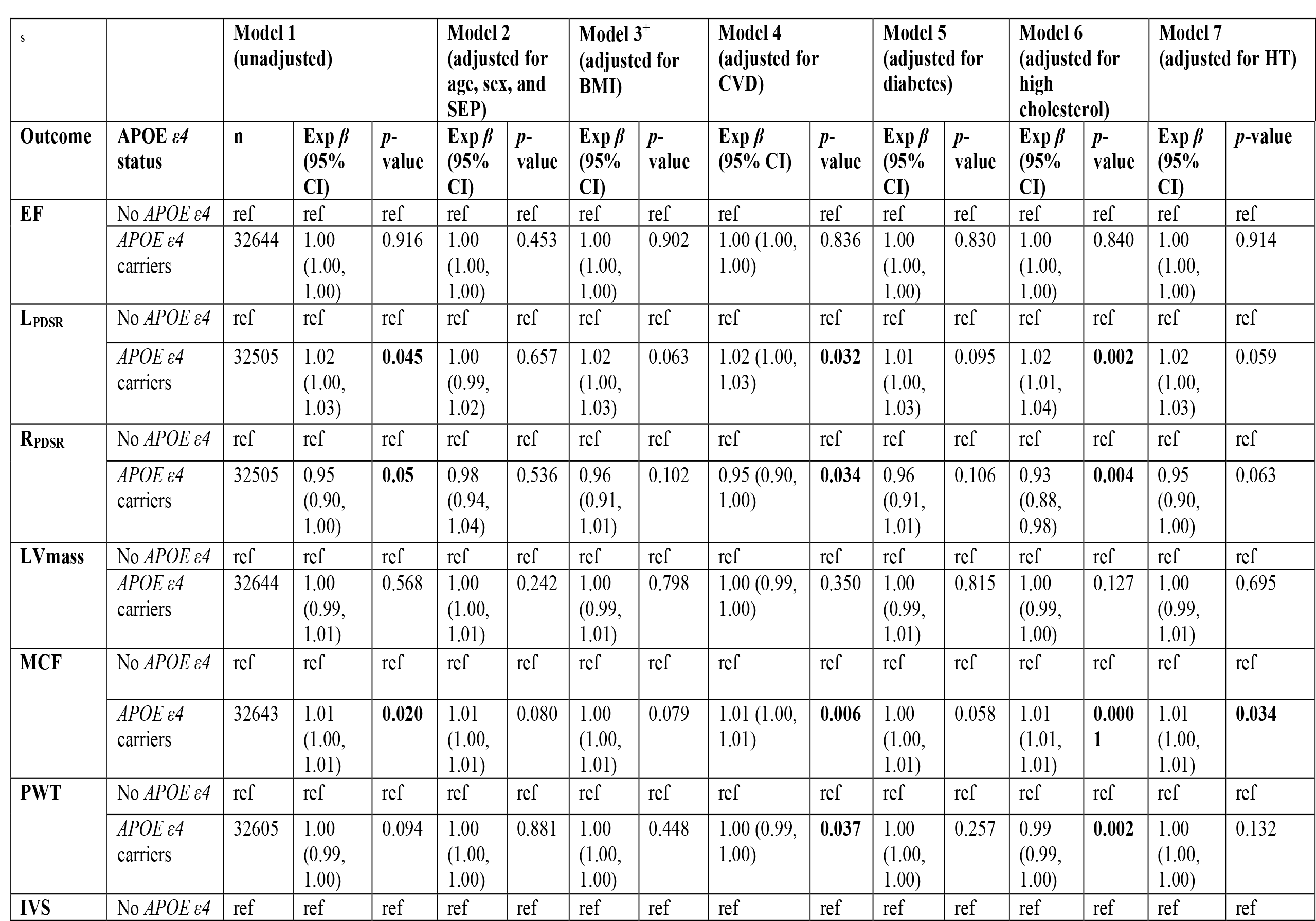
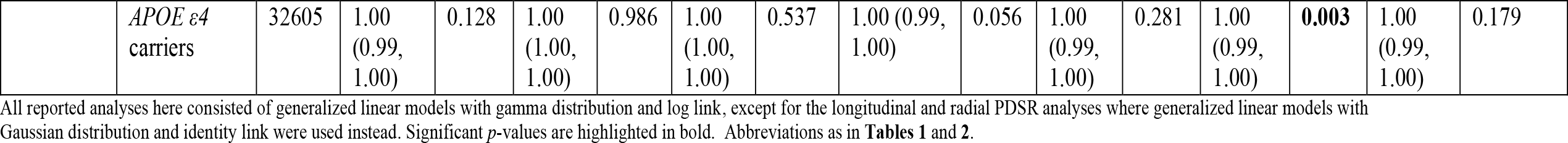
Associations between *APOE ε4* genotypes and echocardiographic data in older age by comparing non-*APOE ε4* (*ε2ε2, ε2ε3, ε2ε3*) with *APOE ε4* (*ε2ε4, ε3ε4 and ε4ε4*) genotypes in UK Biobank.

**Table 4.** Dose response of *APOE ε4* carriage when assessing the association between *APOE ε4* genotype and echocardiographic and CMR data in older age.

|  |  |  | Model 1<br>(unadjusted) |  |  | Model 2<br>(adjusted for<br>age, sex and<br>SEP) |  | Model 3<br>(adjusted for<br>BMI) |  | Model 4<br>(adjusted for<br>CVD) |  | Model 5<br>(adjusted for<br>diabetes) |  | Model 6<br>(adjusted for<br>high<br>cholesterol) |  | Model 7<br>(adjusted for<br>HT) |  |
| --- | --- | --- | --- | --- | --- | --- | --- | --- | --- | --- | --- | --- | --- | --- | --- | --- | --- |
| Outcome: | Cohort | Analysis | n | Exp $\beta$<br>(95%<br>CI) | p-value | Exp $\beta$<br>(95%<br>CI) | p-value | Exp $\beta$<br>(95%<br>CI) | p-value | Exp $\beta$<br>(95%<br>CI) | p-value | Exp $\beta$<br>(95%<br>CI) | p-value | Exp $\beta$<br>(95%<br>CI) | p-value | Exp $\beta$<br>(95%<br>CI) | p-value |
| MCF | UK<br>biobank | *APOE* $\epsilon 4$-<br>linear | 32644 | 1.01<br>(1.00,<br>1.02) | 0.082 | 1.01<br>(1.00,<br>1.02) | 0.152 | 1.01<br>(1.00,<br>1.02) | 0.239 | 1.01<br>(1.00,<br>1.02) | **0.036** | 1.01<br>(1.00,<br>1.02) | 0.123 | 1.01<br>(1.00,<br>1.02) | **0.006** | 1.01<br>(1.00,<br>1.02) | 0.109 |
| | SABRE+<br>NSHD<br>meta-<br>analysis | *APOE* $\epsilon 4$-<br>linear | 2074 | 1.02<br>(0.96,<br>1.08) | 0.544 | 1.02<br>(0.96,<br>1.08) | 0.516 | 1.03<br>(0.97,<br>1.09) | 0.906 | 1.01<br>(0.95,<br>1.07) | 0.729 | 1.00<br>(0.95,<br>1.07) | 0.870 | 1.01<br>(0.94,<br>1.08) | 0.780 | 1.01<br>(0.96,<br>1.07) | 0.670 |
| | UK<br>biobank | *APOE* $\epsilon 4$ -<br>quadratic | 32644 | 1.00<br>(0.99,<br>1.01) | 0.770 | 1.00<br>(1.00,<br>1.01) | 0.424 | 1.00<br>(0.99,<br>1.01) | 0.972 | 1.00<br>(1.00,<br>1.01) | 0.687 | 1.00<br>(1.00,<br>1.01) | 0.711 | 1.00<br>(1.00,<br>1.01) | 0.723 | 1.00<br>(0.99,<br>1.01) | 0.803 |
| | SABRE+<br>NSHD<br>meta-<br>analysis | *APOE* $\epsilon 4$ -<br>quadratic | 2074 | 0.98<br>(0.93,<br>1.03) | 0.475 | 0.98<br>(0.93,<br>1.03) | 0.451 | 0.99<br>(0.94,<br>1.04) | 0.675 | 0.98<br>(0.94,<br>1.02) | 0.327 | 0.98<br>(0.94,<br>1.02) | 0.251 | 0.97<br>(0.92,<br>1.01) | 0.174 | 0.98<br>(0.94,<br>1.02) | 0.312 |
| Longitu-<br>dinal<br>PDSR | UK<br>biobank | *APOE* $\epsilon 4$-<br>linear | 32505 | 1.01<br>(0.98,<br>1.04) | 0.557 | 1.00<br>(0.97,<br>1.03) | 0.855 | 1.01<br>(0.97,<br>1.04) | 0.756 | 1.01<br>(0.98,<br>1.04) | 0.476 | 1.01<br>(0.98,<br>1.04) | 0.678 | 1.02<br>(0.99,<br>1.05) | 0.199 | 1.01<br>(0.97,<br>1.04) | 0.630 |
| | UK<br>biobank | *APOE* $\epsilon 4$ -<br>quadratic | | 0.99<br>(0.97,<br>1.01) | 0.499 | 1.00<br>(0.98,<br>1.02) | 0.640 | 0.99<br>(0.97,<br>1.01) | 0.388 | 0.99<br>(0.97,<br>1.02) | 0.524 | 0.99<br>(0.97,<br>1.02) | 0.524 | 0.99<br>(0.97,<br>1.02) | 0.526 | 0.99<br>(0.97,<br>1.01) | 0.477 |
| Radial<br>PDSR | UK<br>biobank | *APOE* $\epsilon 4$-<br>linear | 32505 | 0.98<br>(0.88,<br>1.09) | 0.725 | 1.02<br>(0.92,<br>1.13) | 0.786 | 0.99<br>(0.89,<br>1.10) | 0.282 | 0.97<br>(0.87,<br>1.08) | 0.614 | 0.99<br>(0.89,<br>1.10) | 0.858 | 0.95<br>(0.85,<br>1.06) | 0.321 | 0.99<br>(0.88,<br>1.10) | 0.789 |
| | UK<br>biobank | *APOE* $\epsilon 4$ -<br>quadratic | | 1.03<br>(0.96,<br>1.11) | 0.379 | 1.02<br>(0.96,<br>1.10) | 0.503 | 1.03<br>(0.96,<br>1.11) | 0.416 | 1.03<br>(0.96,<br>1.11) | 0.405 | 1.03<br>(0.96,<br>1.11) | 0.400 | 1.03<br>(0.96,<br>1.11) | 0.399 | 1.04<br>(0.96,<br>1.11) | 0.365 |

**Table 5.**
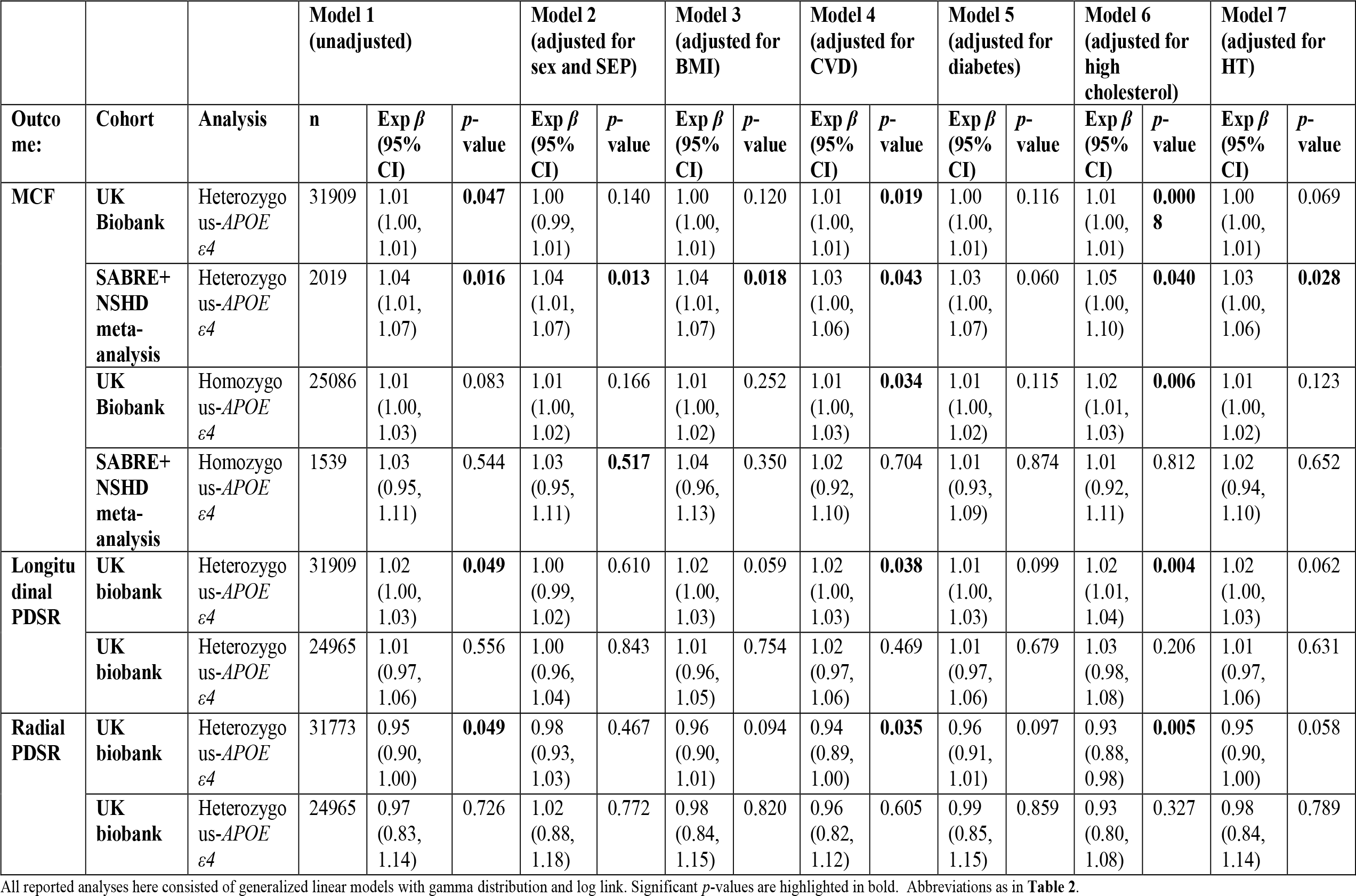
Associations between *APOE ε4* genotypes and echocardiographic data in older age by comparing non-*APOE ε4* (*ε2ε2, ε2ε3, ε2ε3*) with heterozygous-*APOE ε4* (*ε2ε4* and *ε3ε4*) and homozygous-*APOE ε4* (*ε4ε4*) genotypes.

In addition, *APOE ε*4 carriers had a 2% higher longitudinal PDSR (95% CI 0-3%, *p*=0.045), which persisted after adjusting for CVD and diabetes, but was attenuated to 0% in Model 2 and to 1% after adjusting for diabetes (Model 5). Conversely, they had a 5% lower radial PDSR (95% CI 0.90-1.00, *p*=0.05) which behaved similar to longitudinal PDSR on adjustment (**Table 3**).

## DISCUSSION

Data from >35,000 British older adults show that *APOE ε*4 carriage associates with slightly advantageous myocardial performance manifesting as higher MCF and longitudinal strain rates, but slightly lower radial strain rates. A graphical abstract of this work is presented in **Figure 2**. *APOE ε*4 might be another example of antagonistic pleiotropy^6^ as *ε*4 carriage appears to be both beneficial (e.g., fertility and resistance to infections^7^) and detrimental (e.g., Alzheimer’s disease) to human health. The occurrence of the latter further down the fertility timeline in older age might explain the allele’s persistence in spite of natural selection.

**Figure 2.**
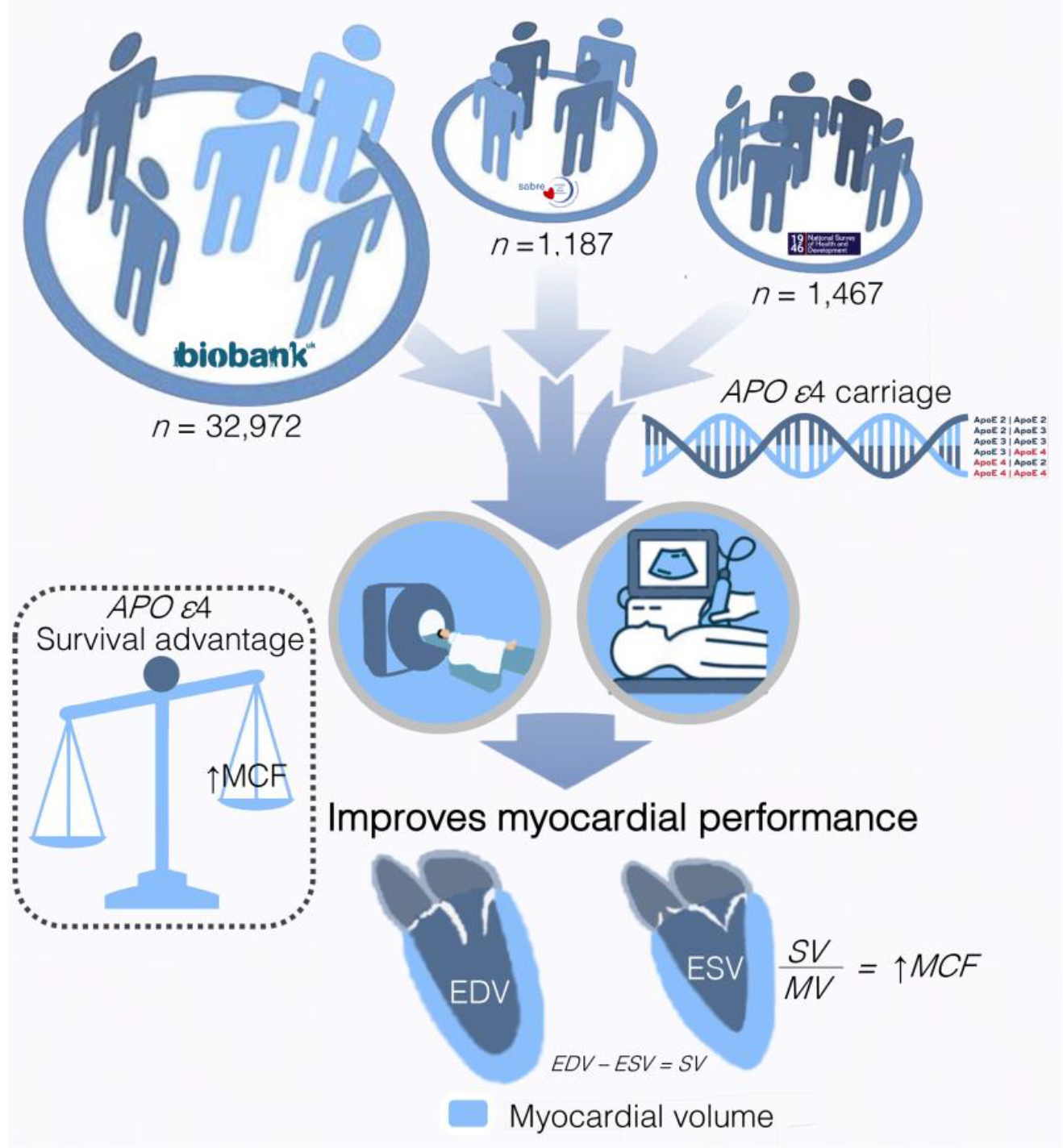
Graphical abstract. Combining data from three British cohorts–1946 National Survey of Health and Development (NSHD), Southall and Brent Revised (SABRE) and UK Biobank–we explored whether *APOE ε*4 carriage associates with beneficial or unfavorable left ventricular (LV) structural and functional parameters by echocardiography and cardiovascular magnetic resonance (CMR) in older age. Based on the presence of *APOE ε*4, genotypes were divided into: *APOE ε*4 (ε2ε4, ε3ε4, *ε*4*ε*4) and non-*APOE ε*4 carriers. Compared to the non-*APOE ε*4 group, *APOE ε*4 carriers had a higher myocardial contraction fraction resulting in greater LV stroke volume generation per 1 mL of myocardium and better longitudinal strain rates compared to non *APOE ε*4 carriers.

In terms of cardiovascular health, *APOE ε*4 carriage was previously associated with CVD (IHD^14^ and myocardial infarction^32^) and CVD risk factors (such as hypertension^12^ and diabetes^13^). Although the exact mechanism is yet to be elucidated, it is postulated that *APOE ε*4 might contribute to the development of metabolic syndrome^33^. *APOE ε*4 differs from *APOE ε*3 at amino acid position 112 where arginine (positively charged side chain) is present instead of cysteine (non-polar side chain). Given its ability to bind to peripheral and hepatic lipoprotein receptors, it is plausible for the *APOE ε* isoforms to have different binding affinities explaining the link with dyslipidemia^14^. However, emerging evidence points to more a complex mechanism as *APOE ε* can also alter the levels of *APOB*^34^ which is itself also associated with CVD^35^. In addition, *APOE ε* is mainly produced by the liver, but can also be synthesized in and regulate the activity of adipocytes^36^ which might explain the relationship between *APOE ε4* and insulin resistance ^33,37^. Here we show that *APOE ε*4 carriage appears to associate with a higher MCF. The MCF is a volumetric index of LV myocardial shortening which captures maladaptive myocardial hypertrophy otherwise missed by conventional biomarkers such as EF, mass, and wall thickness, as it considers the relationship between LVmass and SV^38^. It has been previously associated with CV morbidity and mortality independent of conventional risk factors^39^. In addition, it is regarded as a highly-sensitive metric of systolic function, and low values have been linked to negative outcomes even in the presence of apparently normal LV EF^40^ indicating its strength as a subclinical disease marker. A higher MCF in the context of *APOE ε*4 carriage might mean a slightly advantageous cardiac phenotype in terms of heart function. Dissociable effects of *APOE ε*4 carriage have been previously reported in the context of better attention despite the higher risk of Alzheimer’s disease^10^. Although the literature is sparse, *APOE ε*4 carriage has been previously linked to higher levels of androgens^41^ or dysregulated glucose and ketone metabolism^7^ which could putatively increase myocardial contractility leading to a higher stroke volume per unit of LV mass which is being captured by the MCF^42^ We go on to show that there is likely to be a dose response relationship based on the number of *ε*4 alleles carried by an individual, as per the polynomial contrasts analyses (in the sensitivity analysis the association of homozygous *APOE ε*4 with MCF likely did not persist as only 2.29% of individuals (816) were homozygous for *APOE ε*4). Another explanation is that healthier *APOE ε*4 carriers may have been more likely to survive and/or to participate in the studies resulting in selection bias. This would fit with the known effects of *APOE ε*4 carriage on IHD, HT, lipids, and cognitive function. Previous studies have described cognitive advantages in heterozygotes that were not replicated in the homozygotes^43^ mirroring our data. Indeed, *APOE ε*4 carriage was associated with a greater longitudinal but lower radial strain suggesting that different myocardial contraction dynamics might be contributing to the observed association with MCF. The observed trend linking *APOE ε*4 carriage with slightly better echocardiographic LV filling pressures (lower E/e’ may suggest less ventricular stiffness in some but not all cases^44^), albeit attenuated in multivariable models, lends plausibility to this theory. The CMR analyses indicated a slight association between *APOE ε*4 carriage and thinner ventricular walls, and similarly the echocardiographic analyses found no association between *APOE ε*4 carriage and LV hypertrophy biomarkers (LVPWT_s/d_, IVS_s/d_, LVmass). These data collectively suggest that the observed MCF enhancement is not mediated by pathological ventricular thickening but through improved myocardial energetics and contractility, with calcium potentially implicated^41,42^.

The main strength of our study is that we were able to replicate the findings in three independent cohorts encompassing >35000 individuals, across imaging modalities (echocardiography and CMR). In addition, as the MRC NSHD is a birth cohort, the participants were implicitity age-matched across all the analyses, exposed to similar epoch-related risk factors and had access to similar treatment facilities across the decades. Since both SABRE and NSHD were longitudinal cohorts in which timing of genotyping and echocardiography were not necessarily contemporaneous, selective follow-up may have potentially excluded homozygous or heterozygous individuals who already passed away with the worst cardiac phenotypes.

## CONCLUSION

*APOE ε*4 carriage associates with improved myocardial performance in older age resulting in greater LV stroke volume generation per 1 mL of myocardium and better longitudinal strain rates compared to non *APOE ε*4 carriers. This potentially favorable cardiac phenotype adds to the growing number of reported survival advantages attributed to *APOE* ε4 carriage that might collectively explain its persistence in humans.

## Data Availability

https://www.sabrestudy.org/

https://www.ukbiobank.ac.uk/

## DECLARATIONS

## CONTRIBUTORS’ STATEMENT

G Captur and C Topriceanu conceptualized the study design and implementation, analyzed the data, interpreted the results, and wrote the manuscript. M.Shah, M Webber, F Chan, JC Moon, AD Hughes, N Chaturvedi, J Schott, M Richards, and DP O’Regan were involved in data acquisition and critically reviewed and revised the manuscript. All authors were involved in critically reviewing and revising the manuscript, approved the final version as submitted and agree to be accountable for all aspects of the work.

## REFERENCES

1 Mahley, R. W. Apolipoprotein E: Cholesterol Transport Protein with Expanding Role in Cell Biology. Science (American Association for the Advancement of Science) 240, 622–630 (1988). https://doi.org:10.1126/science.3283935

2 Yamazaki, Y., Zhao, N., Caulfield, T. R., Liu, C.-C. & Bu, G. Apolipoprotein E and Alzheimer disease: pathobiology and targeting strategies. Nature reviews. Neurology 15, 501–518 (2019). https://doi.org:10.1038/s41582-019-0228-7

3 Neu, S. C. et al. Apolipoprotein E Genotype and Sex Risk Factors for Alzheimer Disease: A Meta-analysis. JAMA neurology 74, 1178–1189 (2017). https://doi.org:10.1001/jamaneurol.2017.2188

4 Skoog, I. et al. Association between APOE Genotype and Change in Physical Function in a Population-Based Swedish Cohort of Older Individuals Followed Over Four Years. (2016).

5 Nao, J. et al. Adverse effects of the apolipoprotein E ε4 allele on episodic memory, task switching and gray matter volume in healthy young adults. Frontiers in human neuroscience 11, 346–346 (2017). https://doi.org:10.3389/fnhum.2017.00346

6 Tuminello, E. R. & Han, S. D. The Apolipoprotein E Antagonistic Pleiotropy Hypothesis: Review and Recommendations. International journal of alzheimer’s disease 2011, 726197–726112 (2011). https://doi.org:10.4061/2011/726197

7 Smith, C. J., Ashford, J. W. & Perfetti, T. A. Putative Survival Advantages in Young Apolipoprotein ?4 Carriers are Associated with Increased Neural Stress. Journal of Alzheimer’s disease 68, 885–923 (2019). https://doi.org:10.3233/JAD-181089

8 Jasienska, G. et al. Apolipoprotein E (ApoE) polymorphism is related to differences in potential fertility in women: A case of antagonistic pleiotropy? Proceedings of the Royal Society. B, Biological sciences 282, 20142395–20142395 (2015). https://doi.org:10.1098/rspb.2014.2395

9 Lumsden, A. L., Mulugeta, A., Zhou, A. & Hyppönen, E. Apolipoprotein E (APOE) genotype-associated disease risks: a phenome-wide, registry-based, case-control study utilising the UK Biobank. EBioMedicine 59, 102954–102954 (2020). https://doi.org:10.1016/j.ebiom.2020.102954

10 Wolk, D. A. & Dickerson, B. C. Apolipoprotein E (APOE) genotype has dissociable effects on memory and attentional—executive network function in Alzheimer’s disease. Proceedings of the National Academy of Sciences - PNAS 107, 10256–10261 (2010). https://doi.org:10.1073/pnas.1001412107

11 Zhao, Q. R., Lei, Y. Y., Li, J., Jiang, N. & Shi, J. P. Association between apolipoprotein E polymorphisms and premature coronary artery disease: a meta-analysis. CLIN CHEM LAB MED 55, 284–298 (2017). https://doi.org:10.1515/cclm-2016-0145

12 Shi, J. et al. Association between ApoE polymorphism and hypertension: A meta-analysis of 28 studies including 5898 cases and 7518 controls. GENE 675, 197–207 (2018). https://doi.org:10.1016/j.gene.2018.06.097

13 Chen, D. W., Shi, J. K., Li, Y., Yang, Y. & Ren, S. P. Association between ApoE Polymorphism and Type 2 Diabetes: A Meta-Analysis of 59 Studies. BIOMED ENVIRON SCI 32, 823–838 (2019). https://doi.org:10.3967/bes2019.104

14 Khan, T. A. et al. Apolipoprotein E genotype, cardiovascular biomarkers and risk of stroke: Systematic review and meta-analysis of 14 015 stroke cases and pooled analysis of primary biomarker data from up to 60 883 individuals. INT J EPIDEMIOL 42, 475–492 (2013). https://doi.org:10.1093/ije/dyt034

15 Mueller, K. et al. Brain Damage With Heart Failure: Cardiac Biomarker Alterations and Gray Matter Decline. Circulation research 126, 750–764 (2020). https://doi.org:10.1161/CIRCRESAHA.119.315813

16 Kuh, D. et al. Cohort profile: updating the cohort profile for the MRC National Survey of Health and Development: a new clinic-based data collection for ageing research. Int J Epidemiol, 40 (1) e1–e9. (2011) (2011).

17 Tillin, T., Forouhi, N. G., McKeigue, P. M. & Chaturvedi, N. Southall And Brent REvisited: Cohort profile of SABRE, a UK population-based comparison of cardiovascular disease and diabetes in people of European, Indian Asian and African Caribbean origins. International Journal of Epidemiology 41, 33–42 (2012). https://doi.org:10.1093/ije/dyq175

18 Sudlow, C. et al. UK Biobank: An Open Access Resource for Identifying the Causes of a Wide Range of Complex Diseases of Middle and Old Age. PLoS medicine 12, e1001779–e1001779 (2015). https://doi.org:10.1371/journal.pmed.1001779

19 Kuh, D. et al. Cohort profile: updating the cohort profile for the MRC National Survey of Health and Development: a new clinic-based data collection for ageing research. Int J Epidemiol, 40 (1) e1–e9. (2011) (2011).

20 Al Saikhan, L. et al. Relationship Between Image Quality and Bias in 3D Echocardiographic Measures: Data From the SABRE (Southall and Brent Revisited) Study. Journal of the American Heart Association 11, e019183–e019183 (2022). https://doi.org:10.1161/JAHA.120.019183

21 Chirinos, J. A. et al. Left Ventricular Mass: Allometric Scaling, Normative Values, Effect of Obesity, and Prognostic Performance. Hypertension (Dallas, Tex. 1979) 56, 91–98 (2010). https://doi.org:10.1161/HYPERTENSIONAHA.110.150250

22 Petersen, S. E. et al. UK Biobank’s cardiovascular magnetic resonance protocol. Journal of cardiovascular magnetic resonance 18, 8–8 (2016). https://doi.org:10.1186/s12968-016-0227-4

23 Bai, W. et al. Automated cardiovascular magnetic resonance image analysis with fully convolutional networks. Journal of cardiovascular magnetic resonance 20, 65–65 (2018). https://doi.org:10.1186/s12968-018-0471-x

24 Schulz-Menger, J. et al. Standardized image interpretation and post-processing in cardiovascular magnetic resonance - 2020 update: Society for Cardiovascular Magnetic Resonance (SCMR): Board of Trustees Task Force on Standardized Post-Processing. Journal of cardiovascular magnetic resonance 22, 19–19 (2020). https://doi.org:10.1186/s12968-020-00610-6

25 Cerqueira, M. D. et al. Standardized myocardial segmentation and nomenclature for tomographic imaging of the heart - A statement for healthcare professionals from the Cardiac Imaging Committee of the Council on Clinical Cardiology of the American Heart Association. Circulation (New York, N.Y.) 105, 539–542 (2002). https://doi.org:10.1161/hc0402.102975

26 Thanaj, M. et al. Genetic and environmental determinants of diastolic heart function. (2022).

27 Rousseau, K. et al. MUC7 haplotype analysis: Results from a longitudinal birth cohort support protective effect of the MUC7 5 allele on respiratory function. Annals of human genetics 70, 417–427 (2006). https://doi.org:10.1111/j.1469-1809.2006.00250.x

28 Rawle, M. et al. Apolipoprotein-E (ApoE) ε4 and cognitive decline over the adult life course. (2018).

29 Bycroft, C. et al. The UK Biobank resource with deep phenotyping and genomic data. Nature (London) 562, 203–209 (2018). https://doi.org:10.1038/s41586-018-0579-z

30 Townsend, P., Beattie, A. & Phillimore, P. Health and deprivation : inequality and the north / Peter Townsend, Peter Phillimore and Alastair Beattie. (Routledge, 1989).

31 71–117 (2014).

32 Xu, H. et al. Meta-analysis of apolipoprotein e gene polymorphism and susceptibility of myocardial infarction. PLOS ONE 9, e104608–e104608 (2014). https://doi.org:10.1371/journal.pone.0104608

33 Torres-Perez, E., Ledesma, M., Garcia-Sobreviela, M. P., Leon-Latre, M. & Arbones-Mainar, J. M. Apolipoprotein E4 association with metabolic syndrome depends on body fatness. ATHEROSCLEROSIS 245, 35–42 (2015). https://doi.org:10.1016/j.atherosclerosis.2015.11.029

34 Griffin, B. A. et al. APOE4 Genotype Exerts Greater Benefit in Lowering Plasma Cholesterol and Apolipoprotein B than Wild Type (E3/E3), after Replacement of Dietary Saturated Fats with Low Glycaemic Index Carbohydrates. NUTRIENTS 10, 1524 (2018). https://doi.org:10.3390/nu10101524

35 Richardson, T. G. et al. Evaluating the relationship between circulating lipoprotein lipids and apolipoproteins with risk of coronary heart disease: A multivariable Mendelian randomisation analysis. PLoS medicine 17, e1003062–e1003062 (2020). https://doi.org:10.1371/JOURNAL.PMED.1003062

36 Tejedor, M. T., Garcia-Sobreviela, M. P., Ledesma, M. & Arbones-Mainar, J. M. The apolipoprotein e polymorphism rs7412 associates with body fatness independently of plasma lipids in middle aged men. PLOS ONE 9, e108605–e108605 (2014). https://doi.org:10.1371/journal.pone.0108605

37 Arbones-Mainar, J. M., Johnson, L. A., Altenburg, M. K. & Maeda, N. Differential modulation of diet-induced obesity and adipocyte functionality by human apolipoprotein E3 and E4 in mice. INT J OBESITY 32, 1595–1605 (2008). https://doi.org:10.1038/ijo.2008.143

38 King, D. L., El-Khoury Coffin, L. & Maurer, M. S. Myocardial contraction fraction: a volumetric index of myocardial shortening by freehand three-dimensional echocardiography. Journal of the American College of Cardiology 40, 325–329 (2002). https://doi.org:10.1016/s0735-1097(02)01944-7

39 Chuang, M. L. M. D. et al. Usefulness of the Left Ventricular Myocardial Contraction Fraction in Healthy Men and Women to Predict Cardiovascular Morbidity and Mortality. The American journal of cardiology 109, 1454–1458 (2012). https://doi.org:10.1016/j.amjcard.2012.01.357

40 Rubin, J., Steidley, D. E., Carlsson, M., Ong, M.-L. & Maurer, M. S. Myocardial Contraction Fraction by M-Mode Echocardiography Is Superior to Ejection Fraction in Predicting Mortality in Transthyretin Amyloidosis. Journal of Cardiac Failure 24, 504–511 (2018). https://doi.org:10.1016/j.cardfail.2018.07.001

41 Žofková, I., Zajíčková, K., Hill, M. & Hořínek, A. Apolipoprotein E gene determines serum testosterone and dehydroepiandrosterone levels in postmenopausal women. European journal of endocrinology 147, 503–506 (2002). https://doi.org:10.1530/eje.0.1470503

42 Ayaz, O. & Howlett, S. E. Testosterone modulates cardiac contraction and calcium homeostasis: Cellular and molecular mechanisms. Biology of sex differences 6, 9–9 (2015). https://doi.org:10.1186/s13293-015-0027-9

43 Gharbi-Meliani, A. et al. The association of APOE ε4 with cognitive function over the adult life course and incidence of dementia: 20 years follow-up of the Whitehall II study. (2021).

44 Park, J.-H. & Marwick, T. H. Use and Limitations of E/e’ to Assess Left Ventricular Filling Pressure by Echocardiography. Journal of cardiovascular ultrasound 19, 169 (2011). https://doi.org:10.4250/jcu.2011.19.4.169

