## Supplementary Material for "*APOE ε*4 carriage associates with improved myocardial performance in older age"

---

**Corresponding author:**

Gabriella Captur

Consultant Cardiologist in Inherited Heart Muscle Conditions, Senior Clinical Lecturer

Institute of Cardiovascular Science,

University College London, Gower Street, London WC1E 6BT, UK

**Supplementary Table S1. Associations between *APOE ε4* genotypes and echocardiographic data by comparing non-*APOE ε4* (*ε2ε2, ε2ε3, ε2ε3*) with any *APOE ε4* (*ε2ε4, ε3ε4 and ε4ε4*) genotypes in NSHD.**

| s | | **Model 1 (unadjusted)** | | | **Model 2 (adjusted for sex and SEP)** | | **Model 3+ (adjusted for BMI)** | | **Model 4 (adjusted for CVD)** | | **Model 5 (adjusted for diabetes)** | | **Model 6 (adjusted for high cholesterol)** | | **Model 7 (adjusted for HT)** | |
|---|---|---|---|---|---|---|---|---|---|---|---|---|---|---|---|---|
| **Outcome** | **APOE *ε4* status** | **n** | **Exp *β* (95% CI)** | ***p*-value** | **Exp *β* (95% CI)** | ***p*-value** | **Exp *β* (95% CI)** | ***p*-value** | **Exp *β* (95% CI)** | ***p*-value** | **Exp *β* (95% CI)** | ***p*-value** | **Exp *β* (95% CI)** | ***p*-value** | **Exp *β* (95% CI)** | ***p*-value** |
| **EF** | No *APOE ε4* | ref | ref | ref | ref | ref | ref | ref | ref | ref | ref | ref | ref | ref | ref | ref |
| | *APOE ε4* carriers | 1303 | 0.99 (0.98, 1.01) | 0.421 | 1.00 (0.98, 1.01) | 0.552 | 0.99 (0.98, 1.01) | 0.414 | 1.00 (0.98, 1.01) | 0.524 | 0.99 (0.98, 1.01) | 0.160 | 0.99 (0.98, 1.01) | 0.436 | 1.00 (0.98, 1.01) | 0.524 |
| **E/e'** | No *APOE ε4* | ref | ref | ref | ref | ref | ref | ref | ref | ref | ref | ref | ref | ref | ref | ref |
| | *APOE ε4* carriers | 1342 | 0.98 (0.95, 1.01) | 0.193 | 0.99 (0.96, 1.02) | 0.288 | 0.98 (0.95, 1.01) | 0.186 | 0.97 (0.94, 1.01) | 0.101 | 0.97 (0.94, 1.00) | 0.080 | 0.97 (0.94, 1.01) | 0.115 | 0.98 (0.95, 1.01) | 0.218 |
| **LVmass** | No *APOE ε4* | ref | ref | ref | ref | ref | ref | ref | ref | ref | ref | ref | ref | ref | ref | ref |
| | *APOE ε4* carriers | 1069 | 1.05 (1.00, 1.11) | 0.057 | 1.01 (0.97, 1.06) | 0.665 | 1.05 (1.01, 1.1) | **0.033** | 1.06 (1.00, 1.13) | **0.040** | 1.07 (1.01, 1.13) | **0.024** | 1.03 (0.98, 1.10) | 0.270 | 1.06 (1.00, 1.11) | **0.040** |
| **MCF** | No *APOE ε4* | ref | ref | ref | ref | ref | ref | ref | ref | ref | ref | ref | ref | ref | ref | ref |
| | *APOE ε4* carriers | 916 | 1.06 (1.00,1.12) | **0.050** | 1.06 (1.00, 1.12) | **0.038** | 1.05 (1.00, 1.11) | 0.064 | 1.05 (0.99,1.12) | 0.112 | 1.06 (1.00, 1.12) | 0.056 | 1.08 (1.01, 1.14) | **0.020** | 1.05 (0.99, 1.11) | 0.081 |
| **LVPWTs** | No *APOE ε4* | ref | ref | ref | ref | ref | ref | ref | ref | ref | ref | ref | ref | ref | ref | ref |

| | | | | | | | | | | | | | | | |
|---|---|---|---|---|---|---|---|---|---|---|---|---|---|---|---|
| | *APOE ε4* carriers | 1315 | 1.01 (0.99, 1.04) | 0.207 | 1.01 (0.99, 1.03) | 0.537 | 1.01 (0.99, 1.03) | 0.226 | 1.02 (0.99, 1.04) | 0.152 | 1.02 (1.00, 1.05) | 0.053 | 1.01 (0.99, 1.04) | 0.231 | 1.01 (0.99, 1.04) | 0.190 |
| **$LVPWT_d$** | No *APOE ε4* | ref | ref | ref | ref | ref | ref | ref | ref | ref | ref | ref | ref | ref | ref | ref |
| | *APOE ε4* carriers | 1325 | 1.02 (1.00, 1.05) | 0.076 | 1.01 (0.99, 1.04) | 0.199 | 1.02 (1.00, 1.05) | 0.075 | 1.02 (0.99, 1.05) | 0.125 | 1.02 (0.99, 1.05) | 0.123 | 1.02 (0.99, 1.05) | 0.339 | 1.02 (1.00, 1.05) | 0.066 |
| **$IVS_s$** | No *APOE ε4* | ref | ref | ref | ref | ref | ref | ref | ref | ref | ref | ref | ref | ref | ref | ref |
| | *APOE ε4* carriers | 1317 | 1.00 (0.98, 1.02) | 0.952 | 0.99 (0.97, 1.01) | 0.394 | 1.00 (0.98, 1.02) | 0.877 | 1.01 (0.98, 1.03) | 0.524 | 1.00 (0.98, 1.03) | 0.842 | 0.99 (0.97, 1.01) | 0.432 | 1.00 (0.98, 1.02) | 0.935 |
| **$IVS_d$** | No *APOE ε4* | ref | ref | ref | ref | ref | ref | ref | ref | ref | ref | ref | ref | ref | ref | ref |
| | *APOE ε4* carriers | 1327 | 1.01 (0.98, 1.04) | 0.442 | 1.00 (0.98, 1.03) | 0.834 | 1.01 (0.98, 1.04) | 0.491 | 1.01 (0.98, 1.05) | 0.356 | 1.02 (0.99, 1.05) | 0.239 | 1.00 (0.97, 1.03) | 0.867 | 1.01 (0.98, 1.04) | 0.411 |

All reported analyses here consisted of generalized linear models with gamma distribution and log link. Significant *p*-values are highlighted in bold.

*APOE, apolipoprotein E,* β, beta regression coefficient; *BMI, body mass index;* CI, confidence interval; *EF, ejection fraction;* exp, exponentiated; $IVSs_{/d}$, *interventricular septal thickness in systole/diastole; LVmass, left ventricular mass,* $LVPWT_{s/d}$ *left ventricular posterior wall thickness in systole/diastole;* $MCF_i$, *myocardial contraction fraction; NSHD, National Survey of Health and Development; PDSR, peak diastolic strain rate;* ref, reference.

**Supplementary Table S2. Associations between *APOE ε4* genotypes and echocardiographic data at 60-64 years by comparing non-*APOE ε4* (*ε2ε2, ε2ε3, ε2ε3*) with any *APOE ε4* (*ε2ε4, ε3ε4 and ε4ε4*) genotypes in SABRE.**

| s | | Model 1 (unadjusted) | | | Model 2 (adjusted for age, sex and SEP) | | Model 3[+] (adjusted for BMI) | | Model 4 (adjusted for CVD) | | Model 5 (adjusted for diabetes) | | Model 6 (adjusted for high cholesterol) | | Model 7 (adjusted for HT) | |
|---|---|---|---|---|---|---|---|---|---|---|---|---|---|---|---|---|
| **Outcome** | **APOE *ε4* status** | **n** | **Exp *β* (95% CI)** | ***p*-value** | **Exp *β* (95% CI)** | ***p*-value** | **Exp *β* (95% CI)** | ***p*-value** | **Exp *β* (95% CI)** | ***p*-value** | **Exp *β* (95% CI)** | ***p*-value** | **Exp *β* (95% CI)** | ***p*-value** | **Exp *β* (95% CI)** | ***p*-value** |
| **EF** | No *APOE ε4* | ref | ref | ref | ref | ref | ref | ref | ref | ref | ref | ref | ref | ref | ref | ref |
| | *APOE ε4* carriers | 1160 | 1.02 (1.00, 1.04) | 0.134 | 1.02 (1.00, 1.04) | 0.153 | 1.02 (1.00, 1.04) | 0.144 | 1.01 (0.99, 1.03) | 0.228 | 1.01 (0.99, 1.03) | 0.228 | 1.02 (1.00, 1.05) | 0.109 | 1.02 (1.00, 1.04) | 0.148 |
| **E/e'** | No *APOE ε4* | ref | ref | ref | ref | ref | ref | ref | ref | ref | ref | ref | ref | ref | ref | ref |
| | *APOE ε4* carriers | 1148 | 1.00 (0.96, 1.04) | 0.921 | 1.00 (0.95, 1.04) | 0.894 | 1.01 (1.00, 1.01) | 0.984 | 1.00 (0.96, 1.04) | 0.999 | 1.01 (0.97, 1.06) | 0.651 | 0.99 (0.93, 1.05) | 0.679 | 1.03 (1.01, 1.06) | 0.968 |
| **LVmass** | No *APOE ε4* | ref | ref | ref | ref | ref | ref | ref | ref | ref | ref | ref | ref | ref | ref | ref |
| | *APOE ε4* carriers | 1161 | 1.00 (0.97, 1.04) | 0.918 | 1.01 (0.97, 1.04) | 0.752 | 1.01 (0.97, 1.04) | 0.710 | 1.00 (0.97, 1.04) | 0.829 | 1.01 (0.97, 1.05) | 0.656 | 1.01 (0.96, 1.06) | 0.657 | 1.00 (0.97, 1.04) | 0.846 |
| **MCF** | No *APOE ε4* | ref | ref | ref | ref | ref | ref | ref | ref | ref | ref | ref | ref | ref | ref | ref |
| | *APOE ε4* carriers | 1158 | 1.03 (1.00-1.06) | 0.102 | 1.03 (1.00-1.07) | 0.0961 | 1.03 (1.00-1.07) | 0.084 | 1.02 (0.99, 1.06) | 0.161 | 1.02 (0.99, 1.06) | 0.243 | 1.03 (0.99, 1.07) | 0.211 | 1.03 (0.99, 1.06) | 0.141 |
| **$LVPWT_s$** | No *APOE ε4* | ref | ref | ref | ref | ref | ref | ref | ref | ref | ref | ref | ref | ref | ref | ref |

| | | | | | | | | | | | | | | | |
|---|---|---|---|---|---|---|---|---|---|---|---|---|---|---|---|
| | *APOE ε4* carriers | 1161 | 0.98 (0.96, 1.00) | 0.058 | 0.98 (0.96, 1.00) | 0.067 | 0.98 (0.97, 1.00) | 0.0571 | 0.98 (0.96, 1.00) | **0.046** | 0.98 (0.97, 1.00) | 0.104 | 0.99 (0.96, 1.01) | 0.274 | 0.98 (0.96, 1.00) | 0.065 |
| **$LVPWT_d$** | No *APOE ε4* | ref | ref | ref | ref | ref | ref | ref | ref | ref | ref | ref | ref | ref | ref | ref |
| | *APOE ε4* carriers | 1163 | 0.99 (0.97, 1.01) | 0.268 | 0.99 (0.97, 1.01) | 0.308 | 0.99 (0.97, 1.01) | 0.314 | 0.99 (0.97, 1.01) | 0.289 | 0.99 (0.97, 1.01) | 0.454 | 0.98 (0.95, 1.01) | 0.581 | 0.99 (0.97, 1.01) | 0.293 |
| **$IVS_s$** | No *APOE ε4* | ref | ref | ref | ref | ref | ref | ref | ref | ref | ref | ref | ref | ref | ref | ref |
| | *APOE ε4* carriers | 1161 | 1.00 (0.98, 1.02) | 0.829 | 1.00 (0.98, 1.02) | 0.885 | 1.00 (0.98, 1.02) | 0.934 | 1.00 (0.98, 1.02) | 0.836 | 1.00 (0.98, 1.02) | 0.983 | 1.00 (0.98, 1.03) | 0.946 | 1.00 (0.98, 1.02) | 0.948 |
| **$IVS_d$** | No *APOE ε4* | ref | ref | ref | ref | ref | ref | ref | ref | ref | ref | ref | ref | ref | ref | ref |
| | *APOE ε4* carriers | 1163 | 1.00 (0.97, 1.02) | 0.782 | 1.00 (0.97, 1.02) | 0.822 | 1.00 (0.98, 1.02) | 0.856 | 1.00 (0.98, 1.02) | 0.899 | 1.00 (0.98, 1.03) | 0.862 | 1.00 (0.97, 1.04) | 0.788 | 1.00 (0.98, 1.02) | 0.986 |

All reported analyses here consisted of generalized linear models with gamma distribution and log link. Significant *p*-values are highlighted in bold.

*SABRE, Southall and Brent Revised.* Other abbreviations as in **Supplementary Table S1**.

**Supplementary Table S3. Dose response of *APOE ε4* carriage when assessing the association between *APOE ε4* genotype and echocardiographic data in NSHD.**

| | | | **Model 1 (unadjusted)** | | | **Model 2 (adjusted for age, sex and SEP)** | | **Model 3 (adjusted for BMI)** | | **Model 4 (adjusted for CVD)** | | **Model 5 (adjusted for diabetes)** | | **Model 6 (adjusted for high cholesterol)** | | **Model 7 (adjusted for HT)** | |
|---|---|---|---|---|---|---|---|---|---|---|---|---|---|---|---|---|---|
| **Outcome:** | **Cohort** | **Analysis** | **n** | **Exp *β* (95% CI)** | ***p*-value** | **Exp *β* (95% CI)** | ***p*-value** | **Exp *β* (95% CI)** | ***p*-value** | **Exp *β* (95% CI)** | ***p*-value** | **Exp *β* (95% CI)** | ***p*-value** | **Exp *β* (95% CI)** | ***p*-value** | **Exp *β* (95% CI)** | ***p*-value** |
| **MCF** | **SABRE** | *APOE ε4-linear* | 158 | 1.00 (0.94, 1.08) | 0.901 | 1.01 (0.94, 1.08) | 0.878 | 1.01 (0.94, 1.08) | 0.779 | 1.00 (0.94, 1.07) | 0.974 | 1.00 (0.94, 1.07) | 0.984 | 1.01 (0.93, 1.10) | 0.873 | 1.00 (0.94, 1.07) | 0.985 |
| | **NSHD** | *APOE ε4-linear* | 916 | 1.05 (0.95, 1.17) | 0.354 | 1.05 (0.95, 1.17) | 0.340 | 1.06 (0.96, 1.18) | 0.230 | 1.04 (0.93, 1.17) | 0.525 | 1.02 (0.91, 1.14) | 0.775 | 1.02 (0.91, 1.14) | 0.802 | 1.04 (0.94, 1.16) | 0.452 |
| | **SABRE+ NSHD** | Meta-analysis (linear) | 2074 | 1.02 (0.96, 1.08) | 0.544 | 1.02 (0.96, 1.08) | 0.516 | 1.03 (0.97, 1.09) | 0.906 | 1.01 (0.95, 1.07) | 0.729 | 1.00 (0.95, 1.07) | 0.870 | 1.01 (0.94, 1.08) | 0.780 | 1.01 (0.96, 1.07) | 0.670 |
| | **SABRE** | *APOE ε4* - quadratic | 1158 | 0.98 (0.93, 1.03) | 0.350 | 0.98 (0.93, 1.03) | 0.356 | 0.98 (0.94, 1.03) | 0.404 | 0.98 (0.94, 1.03) | 0.381 | 0.98 (0.94, 1.03) | 0.460 | 0.98 (0.93, 1.04) | 0.499 | 0.98 (0.93, 1.03) | 0.350 |
| | **NSHD** | *APOE ε4* - quadratic | 916 | 0.99 (0.92, 1.06) | 0.713 | 0.98 (0.92, 1.06) | 0.664 | 1.00 (0.93, 1.07) | 0.980 | 0.98 (0.91, 1.07) | 0.660 | 0.96 (0.89, 1.04) | 0.315 | 0.95 (0.88, 1.03) | 0.171 | 0.99 (0.92, 1.06) | 0.676 |
| | **SABRE+ NSHD** | Meta-analysis (quadratic) | 2074 | 0.98 (0.93, 1.03) | 0.475 | 0.98 (0.93, 1.03) | 0.451 | 0.99 (0.94, 1.04) | 0.675 | 0.98 (0.94, 1.02) | 0.327 | 0.98 (0.94, 1.02) | 0.251 | 0.97 (0.92, 1.01) | 0.174 | 0.98 (0.94, 1.02) | 0.312 |

The APOE *ε4* genotypes were coded as an ordered category based on the number of *ε4* possessed. Thus, level 0 encompassed *ε2ε2, ε2ε3, ε2ε3*; level 1 *ε2ε4* and *ε3 ε4*; and level 2 *ε4ε4*. Given the existence of three levels, generalized linear models with gamma distribution and orthogonal polynomial contrasts with 2 equally spaced levels (i.e., linear and quadratic) were employed to look for a dose response by *ε4* variants. Abbreviations as in **Supplementary Tables S1/S2**.

**Supplementary Table S4. Associations between *APOE ε4* genotypes and echocardiographic data at 60-64 years by comparing non-*APOE ε4* (*ε2ε2, ε2ε3, ε2ε3*) with heterozygous-*APOE ε4* (*ε2ε4* and *ε3ε4*) and homozygous-*APOE ε4* (*ε4ε4*) genotypes.**

| | | | **Model 1 (unadjusted)** | | | **Model 2 (adjusted for age, sex and SEP)** | | **Model 3 (adjusted for BMI)** | | **Model 4 (adjusted for CVD)** | | **Model 5 (adjusted for diabetes)** | | **Model 6 (adjusted for high cholesterol)** | | **Model 7 (adjusted for HT)** | |
|---|---|---|---|---|---|---|---|---|---|---|---|---|---|---|---|---|---|
| **Outcome:** | **Cohort** | **Analysis** | **n** | **Exp *β* (95% CI)** | ***p*-value** | **Exp *β* (95% CI)** | ***p*-value** | **Exp *β* (95% CI)** | ***p*-value** | **Exp *β* (95% CI)** | ***p*-value** | **Exp *β* (95% CI)** | ***p*-value** | **Exp *β* (95% CI)** | ***p*-value** | **Exp *β* (95% CI)** | ***p*-value** |
| **MCF** | **SABRE** | Heterozygous-*APOE ε4* | 1129 | 1.03 (1.00, 1.07) | 0.089 | 1.03 (1.00, 1.07) | 0.086 | 1.03 (1.00, 1.07) | 0.079 | 1.03 (0.99, 1.06) | 0.139 | 1.02 (0.99, 1.06) | 0.215 | 1.03 (0.99, 1.08) | 0.201 | 1.03 (0.99, 1.07) | 0.121 |
| | **NSHD** | Heterozygous-*APOE ε4* | 890 | 1.05 (1.00, 1.12) | 0.070 | 1.06 (1.00, 1.12) | 0.053 | 1.05 (0.99, 1.11) | 0.104 | 1.05 (0.99, 1.12) | 0.129 | 1.06 (1.00, 1.13) | **0.049** | 1.08 (1.02, 1.15) | **0.015** | 1.05 (0.99, 1.11) | 0.098 |
| | Combined | Meta-analysis | 2019 | 1.04 (1.01, 1.07) | **0.016** | 1.04 (1.01, 1.07) | **0.013** | 1.04 (1.01, 1.07) | **0.018** | 1.03 (1.00, 1.06) | **0.043** | 1.03 (1.00, 1.07) | 0.060 | 1.05 (1.00, 1.10) | **0.040** | 1.03 (1.00, 1.06) | **0.028** |
| | **SABRE** | Homozygous-*APOE ε4* | 865 | 1.01 (0.92, 1.11) | 0.901 | 1.01 (0.92, 1.11) | 0.882 | 1.02 (0.92, 1.12) | 0.746 | 1.00 (0.91, 1.11) | 0.971 | 1.00 (0.91, 1.10) | 0.980 | 1.01 (0.90, 1.14) | 0.883 | 1.00 (0.91, 1.11) | 0.967 |
| | **NSHD** | Homozygous-*APOE ε4* | 674 | 1.07 (0.93, 1.25) | 0.353 | 1.08 (0.93, 1.25) | 0.336 | 1.09 (0.94, 1.26) | 0.234 | 1.06 (0.90, 1.26) | 0.485 | 1.02 (0.87, 1.21) | 0.786 | 1.02 (0.87, 1.20) | 0.842 | 1.06 (0.92, 1.23) | 0.444 |
| | Combined | Meta-analysis | 1539 | 1.03 (0.95, 1.11) | 0.544 | 1.03 (0.95, 1.11) | 0.517 | 1.04 (0.96, 1.13) | 0.350 | 1.02 (0.92, 1.10) | 0.704 | 1.01 (0.93, 1.09) | 0.874 | 1.01 (0.92, 1.11) | 0.812 | 1.02 (0.94, 1.10) | 0.652 |

All reported analyses here consisted of generalized linear models with gamma distribution and log link. Significant *p*-values are highlighted in bold.

Abbreviations as in Supplementary **Tables S1/S2**.
